# The influence of home versus clinic anal human papillomavirus sampling on high-resolution anoscopy uptake in the Prevent Anal Cancer Self-Swab Study

**DOI:** 10.1101/2023.12.27.23300457

**Authors:** Jenna Nitkowski, Timothy J. Ridolfi, Sarah J. Lundeen, Anna R. Giuliano, Elizabeth Chiao, Maria E. Fernandez, Vanessa Schick, Jennifer S. Smith, Bridgett Brzezinski, Alan G. Nyitray

## Abstract

**Background:** Anal cancer disproportionately affects men who have sex with men (MSM) living with HIV. High-resolution anoscopy (HRA) is an in-clinic procedure to detect precancerous anal lesions and cancer, yet prospective data on factors associated with HRA attendance are lacking. We examined whether anal HPV sampling at home versus in a clinic impacts HRA uptake and assessed HRA acceptability.

**Method:** MSM and trans persons 25 years and older were randomized to home-based self-sampling or clinical sampling. All were asked to attend in-clinic HRA one year later. We regressed HRA attendance on study arm using multivariable Poisson regression and assessed HRA acceptability using *χ*^2^ tests.

**Results:** 62.8% of 196 participants who engaged in screening attended HRA. Although not significant (*p*=0.13), a higher proportion of participants who engaged in clinic-based screening attended HRA (68.5%) compared to home-based participants (57.9%). Overall, HRA uptake was higher among participants with anal cytology history (aRR 1.44, 95% CI 1.11 – 1.87) and lower among participants preferring versatile anal sex position versus insertive (aRR 0.70, 95% CI 0.53 – 0.91), but did not differ by race or HIV serostatus. In the clinic arm, persons living with HIV had lower HRA attendance (42.9%) versus HIV-negative participants (73.3%) (*p*=0.02) and Black non-Hispanic participants had lower HRA attendance (41.7%) than White non-Hispanic participants (73.1%), (*p*=0.04); however, no differences in attendance by race or HIV status were observed in the home arm.

**Conclusions:** HRA uptake differed significantly by race and HIV status in the clinic arm but not the home arm.

## Introduction

Anal cancer disproportionately affects men who have sex with men (MSM) and individuals living with HIV.[1] Over 90% of anal cancers are caused by persistent infection with human papillomavirus (HPV) [2]. Most HPV infections are cleared by the body. However, persistent anal HPV infection can lead to anal high-grade squamous intraepithelial lesions (HSIL) which are discoverable using high-resolution anoscopy (HRA) [3]. Anal HSIL prevalence is highest among MSM living with HIV.[4] Recently the Anal Cancer-HSIL Outcomes Research (ANCHOR) Study, a large clinical trial of persons living with HIV, found that treating anal HSIL was associated with lower risk of progression to anal cancer.[5]

While no official consensus guidelines for anal cancer screening have been released yet, screening for anal cancer in practice typically follows a similar model as cervical cancer screening using cervical Pap cytology. Anal cytology is performed by swabbing the anal canal and if abnormal cells are found, individuals may be referred to HRA (similar to cervical colposcopy) where biopsies are taken from suspicious lesions in order to detect anal HSIL. High-resolution anoscopy is an in-clinic procedure to examine the anal canal. During HRA, a clinician inserts an anoscope into the anal canal and closely examines the anal canal under high magnification. Biopsies may be taken from any areas of concern. Local anesthesia may be injected for biopsies and some patients are prescribed a low-dose anti-anxiety medication prior to the procedure.[6]

Research on HRA attendance and acceptability largely focuses on retrospective studies of clinic populations [7–10]. Prospective studies of HRA attendance, particularly examining factors such as HIV status and race/ethnicity, are limited. Given that persons living with HIV and possibly black MSM disproportionately shoulder the burden of anal cancer [1, 11], research that focuses on these groups and screening uptake is critical.

Our objective in this research was 1) to investigate whether persons who engaged in home-based anal cancer screening had differential uptake of HRA one year later compared to individuals who only engaged in clinic-based anal cancer screening and 2) to assess HRA acceptability. The Prevent Anal Cancer Self-Swab Study was a community-recruited sample of sexual and gender minority individuals aged 25 and older in Milwaukee, Wisconsin who agreed to participate in an anal cancer screening study. We hypothesized that a higher proportion of participants from the clinic-based arm will attend HRA compared to those in the home-based arm. Our rationale is that participants who engaged in clinic screening may be more likely to follow through with other clinic procedures, such as HRA. This study is the first to our knowledge to examine whether anal HPV sampling at home versus in a clinic impacts HRA uptake.

## Methods

### Study recruitment and design

The Prevent Anal Cancer Self-Swab Study recruited MSM and trans persons 25 years and older in the Milwaukee, Wisconsin area to participate in a prospective anal cancer screening study. The protocol for this randomized clinical trial has previously been published.[12] Participants were recruited via social media, flyers, advertisements in local businesses and clinics, and a voluntary referral program. To be eligible for the study, persons must be assigned male sex at birth or identify as transgender, acknowledge sex with men in the last five years or identify as gay or bisexual, be willing to be randomized and able to comply with the protocol, and understand and be willing to give informed consent. Persons who reported use of anticoagulants other than aspirin and non-steroidal anti-inflammatory drugs, reported prior anal cancer diagnosis, planned to move within a year, and reported not be willing to attend one of the designed study clinics at baseline were excluded. All study activities were approved by the Medical College of Wisconsin Human Protections Committee.

Eligible participants were randomized to either a home-based or clinic-based group. Home participants were mailed an anal HPV self-swab kit and clinic participants received anal HPV swabbing from a clinician at their choice of five community clinics. The primary objective of the Prevent Anal Cancer Self-Swab Study was to compare engagement between home-based anal HPV self-sampling and clinic-based sampling.[13] A secondary objective was to examine the influence that these different methods of screening had on HRA uptake, which is the focus of this paper.

Regardless of whether a participant engaged in baseline screening (i.e., home self-sampling or clinician sampling), all participants were asked to attend HRA one year later. There were 10 participants who attended HRA but did not complete baseline screening (Figure 1). Because HPV genotyping is not currently used for anal cancer screening, results from the anal swabs collected at baseline were not provided to participants. However, all participants were asked to receive a digital anal rectal examination (DARE) at the start of the study to check and palpate for abnormalities and/or other symptoms. The DARE was conducted a few weeks after study start for individuals in the home arm so that their engagement in home-based swabbing was not biased by the DARE.

**Fig. 1.**
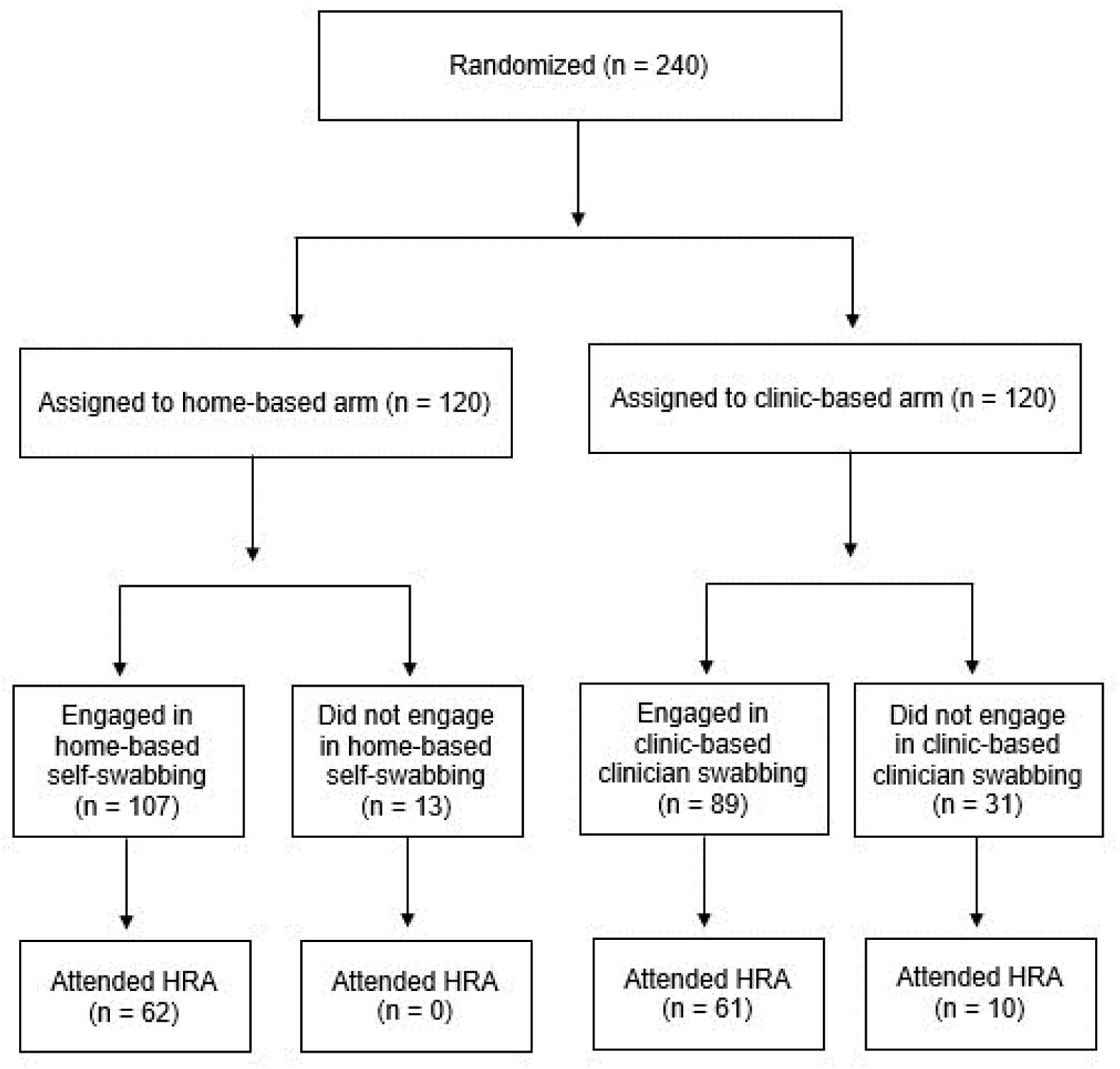
Study flowchart of participants in the Prevent Anal Cancer Self-Swab study, Milwaukee, Wisconsin, 2020-2023

Participants were contacted up to three times via their preferred method of contact to schedule the HRA procedure. High-resolution anoscopy was conducted by a highly trained and experienced high-resolution anoscopist (S.L.) at the Medical College of Wisconsin Anal Dysplasia Program. The clinician documented and biopsied all suspicious lesions in the anal canal or perianal region. In the absence of any suspicious lesions, the anoscopist took two control biopsies from the anal canal to assess for occult HSIL. One person was not biopsied due to starting anti-coagulant use during the study. Immediately after the HRA, participants were asked to complete a computer-assisted self-interview (CASI) which contained questions assessing acceptability of the procedure.

Between 2020 and 2022, a total of 240 participants were randomized to the study. We used participant survey data from those who engaged in baseline screening (n=196) to conduct a per-protocol analysis investigating whether home versus clinic screening, as well as other factors, were associated with HRA uptake. We also performed an intention-to-treat analysis of HRA uptake among all randomized participants in the study (n=240). We then assessed HRA acceptability among those who underwent HRA (n=133).

### Measures

#### Outcome

The outcome was HRA attendance after baseline screening engagement. This was coded as a dichotomous variable indicating whether a participant attended HRA (1=yes, 0=no). Participants could attend HRA up until the end of the study, so there was no time limit on attendance. “Attendance” and “uptake” are used interchangeably throughout the paper for the outcome. Participants who engaged in baseline screening but were withdrawn from the study (n=12) were coded as not attending HRA.

#### Exposures

The primary exposure of interest was engagement in screening. This was a dichotomous variable indicating whether a participant engaged in baseline screening in their respective study arm. For the home arm, this meant returning a mailed home-based anal self-sampling kit. For the clinic arm, this meant making and attending a baseline clinic appointment where a clinician performed anal canal sampling.

Participant demographic and behavioral characteristics as well as screening history and attitudinal items were obtained from the eligibility and baseline surveys. These consisted of age, race/ethnicity, gender identity, sexual orientation, education, HIV status, preferred anal sex position, history of anal cytology, history of HRA, and being afraid of having anal cancer screening for fear of a bad result.

All participants were asked at baseline to attend a clinic to receive a digital anal rectal examination (DARE). Since findings during DARE may impact a participant’s uptake of HRA, we included two measures to examine DARE results. The first is whether an abnormality was detected at either the perianus or anal canal during the DARE, coded as a dichotomous variable (1=yes, 0=no). We also included a variable that captured whether the abnormality identified during the DARE was referred for follow-up (1=yes, 0=no).

Acceptability of HRA was assessed in the post-HRA survey with items guided by Health Belief Model constructs. This included questions about pain, such as *“How much pain did you feel during the HRA?” (nothing at all, a little pain, some pain, a lot of pain, I don’t know)*. Other questions investigated future willingness to undergo HRA, such as “*HRA is something I am willing to do”, “HRA will help me avoid anal cancer”,* and *“After doing HRA, I plan to do an HRA in the future”,* all with response options of strongly agree, agree, disagree, strongly disagree, and I don’t know.

During the time the PAC Study was being conducted, the ANCHOR (Anal Cancer-HSIL Outcomes Research) study released its primary findings. Since the ANCHOR Study indicated that HRA, coupled with treatment of extant HSIL in people with HIV, reduced their risk of anal cancer, we sent a letter to all participants explaining these findings via email on March 31, 2022. The letter contained details about HRA attendance, such as reminders that the procedure could still be scheduled and details about how to schedule the HRA. Since this letter may have affected HRA uptake, we included a variable which assessed whether HRA attendance increased after announcing the ANCHOR Study findings to participants via email using a dichotomous variable (1=attended HRA after the email was sent, 0=attended HRA before the email was sent).

### Statistical analyses

#### Uptake of HRA

Descriptive statistics and Pearson chi-square tests of association were conducted between HRA uptake and participant characteristics. In univariate analysis, likelihood ratio tests were used to test the association between participant characteristics and HRA attendance. Variables with a p-value of less than 0.25 on a likelihood ratio test were included in multivariable regression analyses. We used manual backward elimination to remove exposures with a *p*-value of greater than 0.05 until the remaining exposures had a *p*-value of less than or equal to 0.05. Age, race/ethnicity, education, and HIV were retained in the model as potential confounders. The relative risk was calculated using Poisson regression with robust standard errors and the log-link function in Stata.

#### Acceptability of HRA

The full sample of 133 participants who attended HRA was used to assess HRA acceptability. Descriptive statistics were conducted for the post-HRA survey questions. Second, Pearson chi-square tests were conducted between participant characteristics and reporting HRA pain. Pain during HRA was coded as a dichotomous variable (1=yes, 0=no). Response options of “some pain” or “a lot of pain” were coded as yes, while “no pain” or “a little pain” were coded as no.

All statistical analyses were conducted in SPSS 28(14) and Stata BE 18(15). Fisher’s exact test was used for small cell sizes and all tests were 2-sided.

## Results

The mean age of participants who engaged in baseline screening was 46.4 years and ranged from 25 to 78 years (Table 1). Participants identified as White non-Hispanic (69.7%), Black non-Hispanic (19.0%), Hispanic or Latino/x (10.8%), and Other non-Hispanic (0.5%). Most participants identified as men (95.9%) while 4.1% identified as a trans woman, non-binary, or other gender identity. A total of 86.7% of participants identified as gay and 10.3% identified as bisexual. More than half the sample (64.3%) reported 16 or more years of education. Approximately one in four participants were living with HIV. A total of 7.7% of participants reported having had HRA in the past and 24.0% reported anal cytology in the past.

**Table 1.** Characteristics of participants who engaged in home-based or clinic-based screening stratified by HRA attendance, Prevent Anal Cancer Self-Swab Study, Milwaukee, Wisconsin, 2020-2023, n (%).

|  | TOTAL<br>n=196 | Attended HRA<br>n=123 | Did not attend HRA<br>n=73 |  |
| --- | --- | --- | --- | --- |
|  | n (column %) | n (row %) | n (row %) | p-value |
| **Randomization group** |  |  |  | 0.13 |
| Clinic | 89 (45.4) | 61 (68.5) | 28 (31.5) |  |
| Home | 107 (54.6) | 62 (57.9) | 45 (42.1) |  |
| **Age, years<sup>a</sup> Mean, Range** | 46.4, 25 – 78 | 47.3, 25 – 78 | 45.0, 25 – 74 | 0.27 |
| **Age, years<sup>b</sup>** |  |  |  | 0.40 |
| 25-34 | 58 (29.6) | 35 (60.3) | 23 (39.7) |  |
| 35-44 | 34 (17.3) | 19 (55.9) | 15 (44.1) |  |
| 45-54 | 37 (18.9) | 25 (67.6) | 12 (32.4) |  |
| 55+ | 67 (34.2) | 44 (65.7) | 23 (34.3) |  |
| **Gender identity<sup>c</sup>** |  |  |  | 0.71 |
| Man | 188 (95.9) | 117 (62.2) | 71 (37.8) |  |
| Trans woman, non-binary, or other | 8 (4.1) | 6 (75.0) | 2 (25.0) |  |
| **Sexual orientation<sup>c</sup>** |  |  |  | 0.43 |
| Gay | 169 (86.7) | 108 (63.9) | 61 (36.1) |  |
| Bisexual | 20 (10.3) | 10 (50.0) | 10 (50.0) |  |
| Queer or lesbian | 6 (3.1) | 4 (66.7) | 2 (33.3) |  |
| **Race/ethnicity<sup>c</sup>** |  |  |  | 0.04 |
| White, non-Hispanic | 136 (69.7) | 90 (66.2) | 46 (33.8) |  |
| Black, non-Hispanic | 37 (19.0) | 16 (43.2) | 21 (56.8) |  |
| Hispanic or Latino/x | 21 (10.8) | 15 (71.4) | 6 (28.6) |  |
| Other, non-Hispanic | 1 (0.5) | 1 (100.0) | 0 |  |
| **Education, years<sup>b</sup>** |  |  |  | 0.05 |
| ≤ 12 | 18 (9.2) | 9 (50.0) | 9 (50.0) |  |
| 13-15 | 52 (26.5) | 28 (53.8) | 24 (46.2) |  |
| 16 | 38 (19.4) | 26 (68.4) | 12 (31.6) |  |
| > 16 | 88 (44.9) | 60 (68.2) | 28 (31.8) |  |
| **HIV status** |  |  |  | 0.16 |
| Negative | 148 (75.5) | 97 (65.5) | 51 (34.5) |  |
| Positive | 48 (24.5) | 26 (54.2) | 22 (45.8) |  |
| **Ever had HRA<sup>c</sup>** |  |  |  | 0.18 |
| Yes | 15 (7.7) | 12 (80.0) | 3 (20.0) |  |
| No or don't know | 180 (92.3) | 110 (61.1) | 70 (38.9) |  |
| **Ever had anal cytology** |  |  |  | 0.12 |
| Yes | 47 (24.0) | 34 (72.3) | 13 (27.7) |  |
| No or don't know | 149 (76.0) | 89 (59.7) | 60 (40.3) |  |
| **Preferred anal sex position<sup>c</sup>** |  |  |  | 0.07 |
| Insertive | 41 (21.1) | 30 (73.2) | 11 (26.8) |  |
| Receptive | 54 (27.8) | 38 (70.4) | 16 (29.6) |  |
| Versatile | 94 (48.5) | 50 (53.2) | 44 (46.8) |  |
| Never engaged in anal sex | 5 (2.6) | 3 (60.0) | 2 (40.0) |  |
| **Afraid to have anal cancer screening for fear of a bad result** |  |  |  | 0.08 |
| Strongly agree/agree | 22 (11.3) | 10 (45.5) | 12 (54.5) |  |
| Strongly disagree/disagree | 173 (88.7) | 112 (64.7) | 61 (35.3) |  |
| **Ever been diagnosed with anal warts** |  |  |  | 0.60 |
| Yes | 50 (25.8) | 33 (66.0) | 17 (34.0) |  |
| No | 144 (74.2) | 89 (61.8) | 55 (38.2) |  |
| **Abnormality detected in baseline DARE** |  |  |  | 0.99 |
| Yes | 70 (41.4) | 48 (68.6) | 22 (31.4) |  |
| No | 99 (58.6) | 68 (68.7) | 31 (31.3) |  |
| Did not attend DARE | 27 | 7 | 20 |  |
| **Abnormality referral** |  |  |  | 0.68 |
| Yes | 20 (28.6) | 13 (65.0) | 7 (35.0) |  |
| No | 50 (71.4) | 35 (70.0) | 15 (30.0) |  |

Abbreviations: HRA, high-resolution anoscopy. DARE, digital anal rectal examination. Missing: sexual orientation n=1; race/ethnicity n=1, ever had HRA n=1; preferred anal sex position n=2; afraid to have anal cancer screening for fear of a bad result n=1; ever been diagnosed with anal warts n=2. Pearson chi-square tests were conducted unless otherwise noted.
<sup>a</sup> t-test.
<sup>b</sup> Cochran-Armitage test for trend.
<sup>c</sup> Fisher's exact test.

### Uptake of high-resolution anoscopy

Of the 240 individuals randomized into the home or clinic arm, 133 persons (55.4%) attended HRA (Table 2), 62 (25.8%) in the home arm and 71 (29.6%) in the clinic arm (*p*=0.24).

**Table 2.**
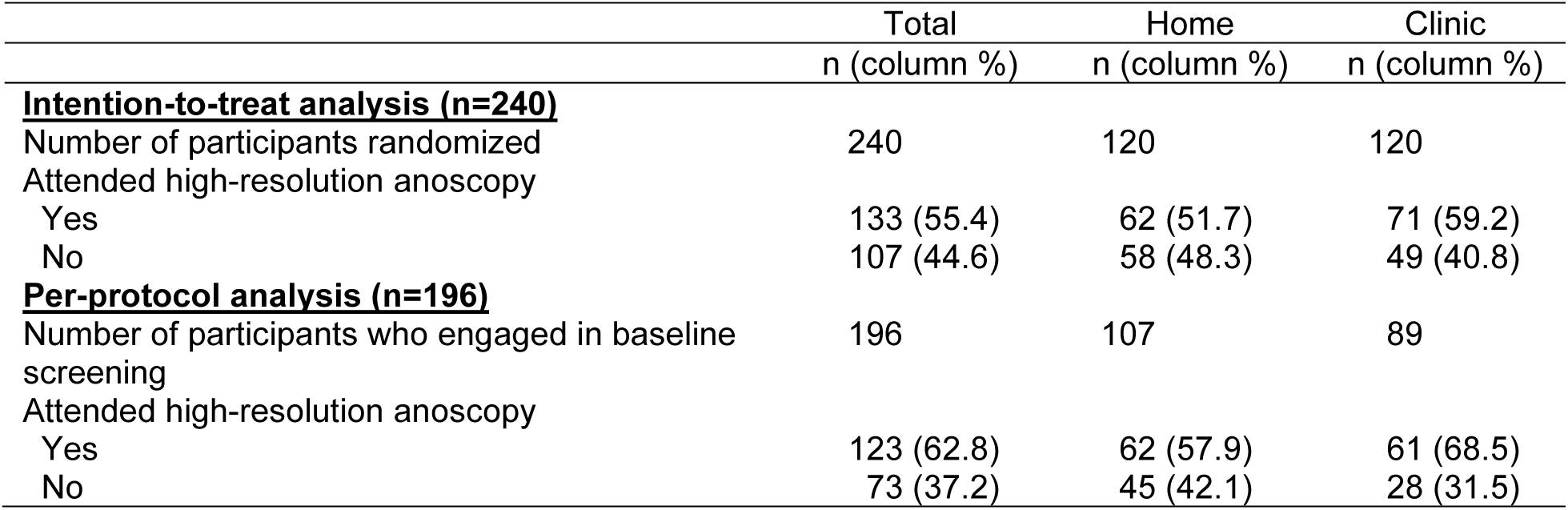
Intention-to-treat and per-protocol analyses of HRA attendance by study arm in the Prevent Anal Cancer Self-Swab Study, Milwaukee, WI, 2020-2023.

Of the 196 participants who engaged in baseline screening, 62.8% attended HRA. Although not significant (*p*=0.13), a higher proportion of clinic-based participants attended HRA (68.5%, n=61) compared to home-based participants (57.9%, n=62) (Table 1). No significant differences in HRA uptake were found by age, gender identity, sexual orientation, both overall and when stratified by study arm (Supplemental Table 1). Overall, Black non-Hispanic participants had the lowest proportion of HRA attendance (43.2%) compared to White non-Hispanic participants (66.2%) and Hispanic participants (71.4%) (*p*=0.04).

In the home arm, 44.0% of Black non-Hispanic participants who engaged in home-based anal self-sampling attended HRA compared to 59.4% of White non-Hispanic participants (*p*=0.18) and 75.0% of Hispanic participants (*p*=0.09) (Figure 2). There was no significant difference in HRA attendance between White non-Hispanic participants and Hispanic participants who engaged in home-based screening (*p*=0.36).

**Fig. 2.**
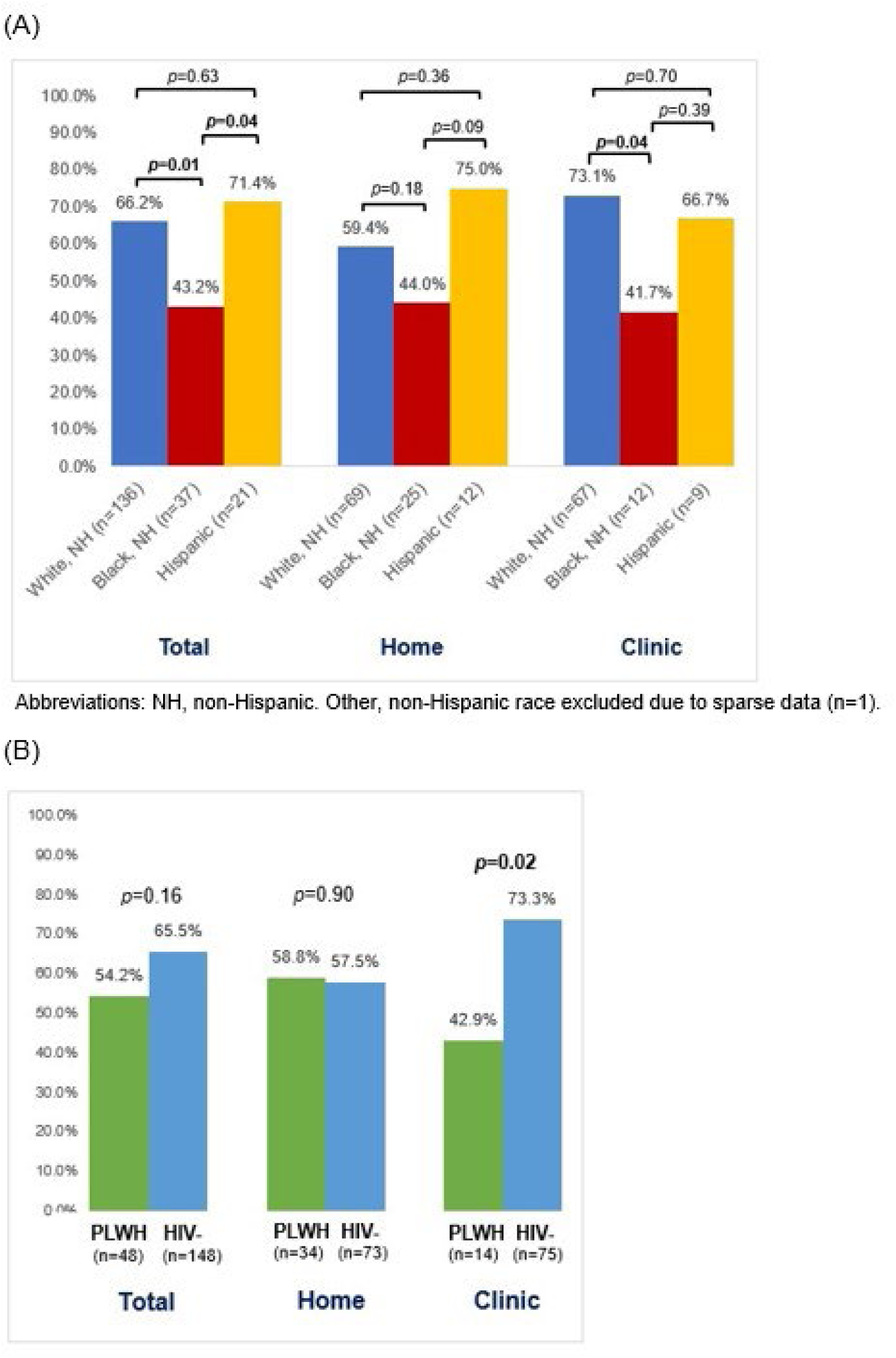
Proportion of participants who engaged in baseline screening and attended high-resolution anoscopy by study arm stratified by (A) race and ethnicity and (B) HIV status in the Prevent Anal Cancer Self-Swab Study, Milwaukee, Wisconsin, 2020-2023 (n=196)

In the clinic arm, Black non-Hispanic participants who engaged in clinic-based screening had a significantly lower proportion of HRA attendance (41.7%) compared to White non-Hispanic participants (73.1%) (*p*=0.04). There was no significant difference in HRA attendance between Black non-Hispanic participants and Hispanic participants (*p*=0.39) or between White non-Hispanic participants and Hispanic participants (*p*=0.70).

Among those who completed home-based anal self-sampling, nearly identical proportions of participants living with HIV (58.8%) and HIV-negative participants (57.5%) attended HRA (*p*=0.90). However, among those in the clinic arm who engaged in baseline screening, a higher proportion of HIV-negative participants attended HRA (73.3%) compared to participants living with HIV (42.9%) (*p*=0.02).

Results of regression analyses showed that ever having anal cytology was significantly associated with HRA attendance after controlling for potential confounders (aRR 1.44, 95% CI 1.11 – 1.87) (Table 3). Participants who reported a versatile anal sex position were less likely to attend HRA compared to those who reported an insertive position (aRR 0.70, 95% CI 0.53 – 0.91). Race had a borderline significant association with HRA attendance, with Black non-Hispanic participants less likely to attend HRA (aRR 0.67, 95% CI 0.44 – 1.01) compared to White non-Hispanic participants.

**Table 3.** Factors associated with HRA attendance among those who engaged in baseline screening in the Prevent Anal Cancer Self-Swab Study, Milwaukee, Wisconsin, 2020-2023, n=196.

|  | Univariate RR (95% CI) | Adjusted RR<sup>a</sup> (95% CI) |
| --- | --- | --- |
| **Randomization group** |  |  |
| Home | 1.0 | 1.0 |
| Clinic | 1.18 (0.95 – 1.47) | 1.17 (0.94 – 1.44) |
| **Age, years** |  |  |
| 25-34 | 1.0 | 1.0 |
| 35-44 | 0.93 (0.64 – 1.33) | 0.96 (0.66 – 1.38) |
| 45-54 | 1.12 (0.82 – 1.52) | 1.12 (0.82 – 1.53) |
| 55+ | 1.09 (0.83 – 1.43) | 1.12 (0.82 – 1.52) |
| **Race/ethnicity<sup>b</sup>** |  |  |
| White, non-Hispanic | 1.0 | 1.0 |
| Black, non-Hispanic | **0.65 (0.44 – 0.96)** | 0.67 (0.44 – 1.01) |
| Hispanic or Latino/x | 1.08 (0.80 – 1.45) | 1.18 (0.85 – 1.64) |
| **Education** |  |  |
| ≤ 12 years | 1.0 | 1.0 |
| 13-15 years | 1.08 (0.64 – 1.82) | 1.10 (0.64 – 1.88) |
| 16 years | 1.37 (0.82 – 2.28) | 1.19 (0.68 – 2.09) |
| > 16 years | 1.36 (0.84 – 2.21) | 1.18 (0.70 – 2.01) |
| **HIV status** |  |  |
| Negative | 1.0 | 1.0 |
| Positive | 0.83 (0.62 – 1.10) | 0.79 (0.55 – 1.14) |
| **Ever had HRA** |  |  |
| No or don't know | 1.0 | n.s. |
| Yes | 1.31 (0.99 – 1.73) | n.s. |
| **Ever had anal cytology** |  |  |
| No or don't know | 1.0 | 1.0 |
| Yes | 1.21 (0.97 – 1.51) | **1.44 (1.11 – 1.87)** |
| **Preferred anal sex position** |  |  |
| Insertive | 1.0 | 1.0 |
| Receptive | 0.96 (0.75 – 1.24) | 0.95 (0.73 – 1.23) |
| Versatile | **0.73 (0.56 – 0.95)** | **0.70 (0.53 – 0.91)** |
| Never engaged in anal sex | 0.82 (0.39 – 1.72) | 1.00 (0.45 – 2.21) |
| **Afraid to have anal cancer screening for fear of a bad result** |  |  |
| Strongly agree/agree | 0.70 (0.44 – 1.13) | n.s. |
| Strongly disagree/disagree | 1.0 | n.s. |

Abbreviations: HRA, high-resolution anoscopy; RR, relative risk, CI, confidence interval, n.s. = not significant. Bolded variables have 95% CI that do not include unity.
<sup>a</sup> The final multivariable model includes all remaining variables in addition to age, race/ethnicity, education, and HIV status which were forced into the model as potential confounders.
<sup>b</sup> Other, non-Hispanic excluded due to small cell size (n=1).

Finally, we did not find any evidence that the ANCHOR letter impacted HRA attendance. Participants who were notified to schedule their HRA after the ANCHOR letter was sent out were not significantly more likely to attend HRA compared to those who were notified to schedule their HRA before the letter was sent (RR 1.18, 95% CI 0.86 – 1.62).

### Acceptability of high-resolution anoscopy

Of the 133 participants who attended HRA, 51.9% reported a little pain, 15.0% reported some pain, and 3.8% reported a lot of pain during the HRA (Table 4). Age was significantly associated with reporting some or a lot of pain during the HRA. Participants ages 25 to 34 years old represented the highest proportion of those who reported some pain or a lot of pain during the HRA (52.0%) compared to other age groups such as those age 55 years and over (28.0%) (*p*=0.02, results not shown).

**Table 4.** Acceptability of high-resolution anoscopy in the Prevent Anal Cancer Self-Swab Study, Milwaukee, Wisconsin, 2021-2023 (n=133).

|  | n (%) |
| --- | --- |
| **How much pain did you feel during the HRA?** |  |
| Nothing at all | 39 (29.3) |
| A little pain | 69 (51.9) |
| Some pain | 20 (15.0) |
| A lot of pain | 5 (3.8) |
| I don't know | -- |
| **If it was recommended to do HRA, I would probably do it how often?** |  |
| Once a month or less | 4 (3.0) |
| Once every few months | 22 (16.5) |
| Once a year | 98 (73.7) |
| Probably never | 1 (0.8) |
| I don't know | 8 (6.0) |
| **HRA is something I am willing to do** |  |
| Strongly agree | 72 (54.1) |
| Agree | 55 (41.4) |
| Disagree | 1 (0.8) |
| Strongly disagree | 2 (1.5) |
| I don't know | 3 (2.3) |
| **HRA will help me avoid anal cancer** |  |
| Strongly agree | 68 (51.1) |
| Agree | 47 (35.3) |
| Disagree | 5 (3.8) |
| Strongly disagree | 1 (0.8) |
| I don't know | 12 (9.0) |
| **After doing HRA, I plan to do an HRA in the future** |  |
| Strongly agree | 40 (30.3) |
| Agree | 46 (34.8) |
| Disagree | 5 (3.8) |
| Strongly disagree | -- |
| I don't know | 41 (31.1) |
| Missing | 1 |

Abbreviations: HRA, high-resolution anoscopy.

The majority of HRA participants reported that they would do HRA once a year (73.7%) if it was recommended. A total of 95.5% of HRA participants strongly agreed or agreed that HRA is something they are willing to do, and 86.5% agreed with the statement that HRA will help them avoid anal cancer. After doing HRA, 65.2% strongly agreed or agreed that they plan to do HRA in the future.

## Discussion

This study is the first to our knowledge to examine whether anal HPV sampling at home versus in a clinic impacts HRA uptake. Using a community-recruited sample of sexual and gender minority individuals, we were interested in investigating how different screening methods may affect attending an in-person clinic appointment for HRA. Overall, HRA uptake was relatively low and did not significantly differ by study arm. Whether a participant collected an anal self-sample at home or a clinician conducted an anal sample from the participant in a clinic did not impact attendance at HRA, although a higher proportion of clinic participants who engaged in baseline screening attended HRA compared to home participants.

However, important significant differences emerged by HIV status and race among those who engaged in clinic screening. In the clinic arm, PLWH had lower HRA attendance than HIV-negative individuals (*p*=0.02). Black non-Hispanic participants in the clinic arm had lower HRA attendance than White non-Hispanic individuals (*p*=0.04). In both the clinic and home arms, Black non-Hispanic participants had the lowest proportions of HRA attendance compared to other racial/ethnic groups. In a study of anal HSIL follow-up, Silvera et al. (2021) found that Black patients and PLWH were less likely to undergo anal HSIL treatment within six months of their diagnosis.[10] Another study of patients with anal squamous cell carcinoma found that persons living with HIV had higher proportions of not receiving treatment than HIV-negative patients[16].

Lower uptake of a clinic procedure may be due to factors at the individual, provider, and system level, such as past discrimination experienced from health care providers or competing demands on time or resources. Another explanation which may be particularly relevant for PLWH is appointment fatigue due to managing and attending multiple appointments for a chronic illness. This might be why we observed lower HRA uptake among participants reporting a preferred versatile anal sex position compared to insertive, given that insertive participants had the lowest proportions of living with HIV. Among mailed home-based anal HPV self-sampling, on the other hand, significant differences in HRA attendance by HIV status, race/ethnicity, or any other participant characteristic were not observed.

There were 10 clinic-based participants who did not engage in screening but attended HRA. These participants were older (median age of 58 years) and a greater proportion were living with HIV (60%). It is possible that these participants may have had concerns about going to a (voluntary) clinic visit during COVID. In contrast, none of the 13 home-based participants who did not complete baseline screening attended HRA. Furthermore, the overall proportion of home-based participants who attended HRA was lower than that of clinic-based participants. Future research is needed on home-based anal self-sampling and HRA uptake.

High-resolution anoscopy infrastructure in the United States is currently limited, with large geographic variation.[17] Follow-up care, in practice, may therefore not be distributed equally for sexual and gender minority individuals across the U.S. due to where they live and lack of HRA providers. Furthermore, self-reported history of anal cytology was significantly associated with HRA uptake. This suggests that those who already have received anal cancer screening were more likely to attend HRA possibly due to better health care access, or greater comfort and/or familiarity with anal procedures. Note that this sample of sexual and gender minority individuals also willingly decided to participate in this study about anal cancer screening and thus their attitudes may differ from other sexual and gender minority individuals who did not participate. Although we did not find a significant difference in HRA uptake by study arm, it is possible that the sample size may have limited the ability to detect differences.

Other potential limitations concern our post-HRA survey question about pain. The two control biopsies taken during HRA may have increased pain levels for participants. Another potential limitation is that we asked about HRA pain immediately after the procedure was conducted. Other research indicates that HRA pain may be significantly higher days *after* the procedure compared to pain *during* the procedure [8] [18]. Thus, our findings may underreport the level of pain experienced by participants from HRA since they may have experienced greater pain in the hours or days following the procedure. Hillman et al. (2011) asked participants about pain during the HRA as well as pain after the HRA, and found that both were negatively correlated with acceptability of HRA [19]. Another study found that patients with high-grade anal dysplasia preferred to undergo HRA under anesthesia due to discomfort [20]. Given that pain and discomfort are such significant barriers to screening, this is an important area of research to inform anal cancer screening guidelines.

Most participants who attended HRA reported that they would be willing to do HRA once a year if it was recommended. This also carries implications for anal cancer screening. For example, a person discovered to have HSIL during HRA might be asked to return six months later for treatment. If HRA is largely acceptable once a year, this may impact HRA uptake if persons are asked to undergo this procedure more frequently.

## Conclusion

This study is the first to compare the influence that home versus clinic anal HPV sampling has on HRA uptake. Overall, over half of participants attended HRA. Most participants who attended HRA strongly agreed or agreed that they would be willing to do HRA in the future. Reported pain during HRA was higher among younger participants. Although not significant, a higher proportion of clinic participants attended HRA compared to home-based participants. While persons living with HIV and Black non-Hispanic participants in the clinic arm had lower HRA attendance compared to HIV-negative and other racial/ethnic groups, no significant differences in HIV status or race were found in the home arm. Given that PLWH and Black MSM are disproportionately affected by anal cancer, interventions are needed to support their clinic attendance.

## Acknowledgements

Thank you to the study participants, the PAC community advisory board, and the PAC Study Team (Cameron Liebert, Christopher Ajala, Madison Humphry, Esmeralda Lezama-Ruiz, Maritza Pallo, and Lisa Rein). Thanks to the clinicians who participated in this study: Mary Kay Schuknecht, Kathryn Kerhin, Nickie Gerboth, Dave Wenten, Annie Lakatos, Andrew Petroll, Brian Hilgeman, Leslie Cockerham, Sol del Mar Aldrete Audiffred, Winsome Panton, Christine Hogan, and Janaki Shah.

## Declarations

### Data Availability Statement

Fully de-identified datasets and data dictionary will be shared with properly trained investigators on the study website within 1 year of study completion after assessment of institutional policies, Medical College of Wisconsin Human Research Protections Program rules, and local, state, and federal laws and regulations.

### Ethics Approval and Patient Consent

Informed consent was obtained from all study participants and study activities were approved by the Medical College of Wisconsin Human Research Protections Committee (protocol #PRO00032999).

---

**Supplemental Table 1.**
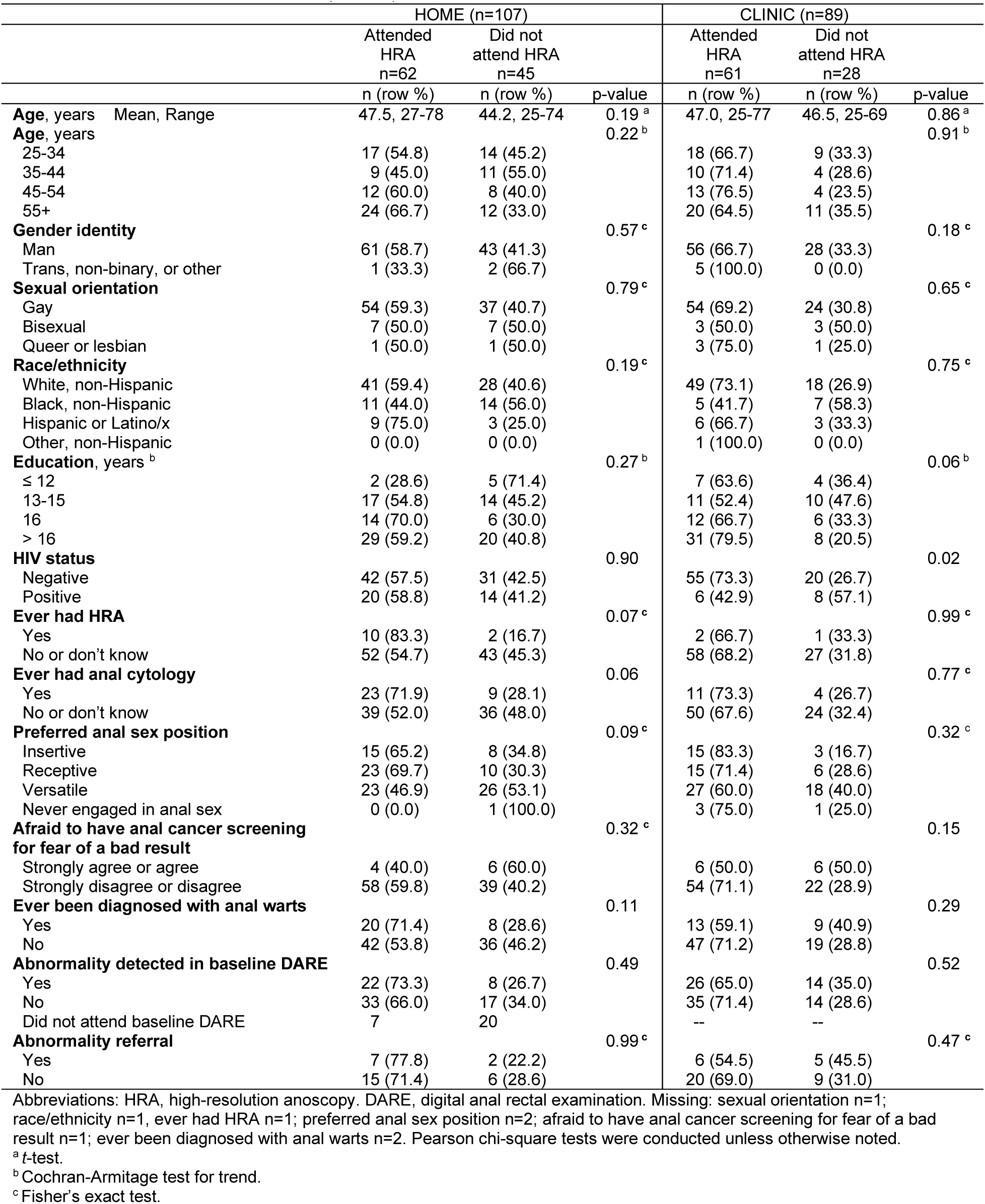
Characteristics of participants who engaged in home-based or clinic-based screening, stratified by study arm and HRA attendance in the Prevent Anal Cancer Self-Swab Study, Milwaukee, Wisconsin, 2020-2023 (n=196).

